# The lived experience of social anxiety disorder: A conceptual model based on published literature and social media listening

**DOI:** 10.1101/2024.11.28.24318127

**Authors:** Ana Lucía Schmidt, Hannah Staunton, Murray B. Stein, Anne Marie Hayes, Raul Rodriguez-Esteban, Kathrin Fischer, Tammy McIver, Eugénie E. Suter

**Author notes:** H Staunton and AL Schmidt contributed equally to this work. Kathrin Fischer has changed affiliation since completing this research. **Corresponding author:** Ana Lucía Schmidt Corresponding author permanent address and contact details: Roche Innovation Center Basel, F. Hoffmann-La Roche Ltd, Grenzacherstrasse 124, 4070 Basel, Switzerland.

## Abstract

Social anxiety disorder (SAD) affects up to 1 in 8 individuals over their lifetime and is characterized by an intense fear of social situations where there may be exposure to unfamiliar people or possible scrutiny. The analysis of social media data rather than traditional methods (interviews or focus groups) can provide a unique opportunity to understand the lived experience of individuals with SAD, for whom interacting with strangers is challenging. This retrospective observational study reviewed published literature from PubMed and data from Reddit using social media listening (SML). A stepwise analysis in line with US Food and Drug Administration Patient-Focused Drug Development guidelines was performed to develop a conceptual model for SAD. Natural language processing techniques and machine learning approaches were employed to extract symptoms and impacts described by individuals with SAD. Eleven publications from the literature and 535,544 posts from 118,040 Reddit users were included. Clinical and patient experts then refined the conceptual model covering three key symptom domains (physical, negative automatic thoughts, and emotions) and two impact domains (social functioning and occupational/educational functioning). This study provides insights into the lived experience of individuals with SAD and confirms the value of SML when traditional methods are inappropriate.

## 1. Introduction

Social anxiety disorder (SAD) affects up to 1 in 8 individuals over their lifetime and represents a substantial burden on individuals and families [1, 2]. SAD is characterized by marked and persistent fear of one or more social situations in which the person is exposed to unfamiliar people or to possible scrutiny by others [2, 3]. The Diagnostic and Statistical Manual of Mental Disorders, Fifth Edition further states “the social situations almost always provoke fear or anxiety,” and anticipatory anxiety may cause avoidance of these situations, interfering significantly with the person’s daily living [3].

The age of onset of SAD is typically in early adolescence, but diagnosis may take longer, with many individuals never being diagnosed [4]. The life-long disease course is often complicated by depression and suicidality [2, 4]. Symptoms (e.g. reflecting inhibited temperament or behavioral inhibition) may be noted early in childhood and increase risk for later SAD diagnosis, but have limited predictive value [5]. While SAD is sometimes diagnosed before the age of 10 years, social anxiety in pre-adolescent children is often accompanied by comorbid disorders that are more easily identified, most commonly other anxiety disorders [6]. Social anxiety in children can also be expressed as selective mutism and specific phobia, making diagnosis difficult at an early age [6-8].

In pediatric individuals (≤18 years), the standard of care (SoC) treatment is cognitive behavioral therapy (CBT) [2, 8]. The current SoC for adults with SAD vary from country to country. SoC guidelines in many countries such as Germany, USA and Japan (among others) recommend a choice of CBT or pharmacological treatment, such as selective serotonin reuptake inhibitors (SSRIs) or serotonin–norepinephrine reuptake inhibitors, as first-line therapy in cases of moderate-to-severe social anxiety [9-11]. However, there is a high unmet need, with a significant proportion of non-responders to CBT or SSRIs in the adult and pediatric populations (40%–60% over 3–5 months of treatment) [12]. Both CBT and pharmacological interventions have a delayed onset of efficacy of several weeks from the initiation of treatment and, additionally, provider availability for CBT and up-titration for SSRIs can cause further delays, while many individuals never receive treatment [2, 8, 13-15].

There is limited information regarding the lived experience of individuals with SAD. One framework that can fill the gap is a conceptual model, which provides a visual depiction of the interrelationships between disease-related symptoms and their impact [16-18]. The US Food and Drug Administration (FDA) guidelines state “a conceptual model can be useful for representing patients’ specific health experiences that result from their disease/condition, the health concepts that describe those specific experiences, and the concept(s) of interest selected for assessment” [18]. Typically, regulators (e.g. the FDA Patient-Focused Drug Development series) recommend speaking directly with affected individuals to ascertain the symptoms and impacts of most relevance [18, 19]. Traditional methods for developing conceptual models include a review of published literature and collecting data directly from individuals with the condition using qualitative methods through different interview types such as one-to-one or group interviews [16, 20]. Previous studies have used social media to complement traditional methods to better understand the patient experience in diseases such as cancer, multiple sclerosis and Parkinson’s disease, and various mental health conditions [21-25]. In SAD, where interacting with strangers is challenging, typical methodological approaches such as interviews and focus groups may not be optimal due to potential distress to participants and potential bias of results (participation and recruitment bias). The review of social media data contributed directly by affected individuals may inform on the lived experience of individuals living with conditions that make social interaction difficult and may be valuable as a primary research source, or as a supplement to traditional methods [26].

The review of social media data in this study provided a unique opportunity to act as a primary research method to limit the burden on individuals with SAD [23, 27, 28]. Thus, this retrospective observational study sought to explore existing qualitative data from the published literature and direct contributions from individuals with SAD on social media sources, to understand the lived experience of SAD in terms of the symptoms experienced and their impact on functioning.

## 2. Materials and methods

A stepwise approach (Fig. 1) was followed in line with Patient-Focused Drug Development FDA guidance [18, 19]. A targeted literature search was used as the basis for creating a preliminary conceptual model for SAD, whereafter social media listening (SML) data and expert feedback were incorporated to yield the final model. Three research questions were studied in the development of the conceptual model:

1. What symptoms are experienced by individuals with SAD?
2. What are the impacts on functioning or daily living for individuals with SAD?
3. Could the conceptual model be used to evaluate clinical outcome assessments used for SAD?

**Fig. 1.**
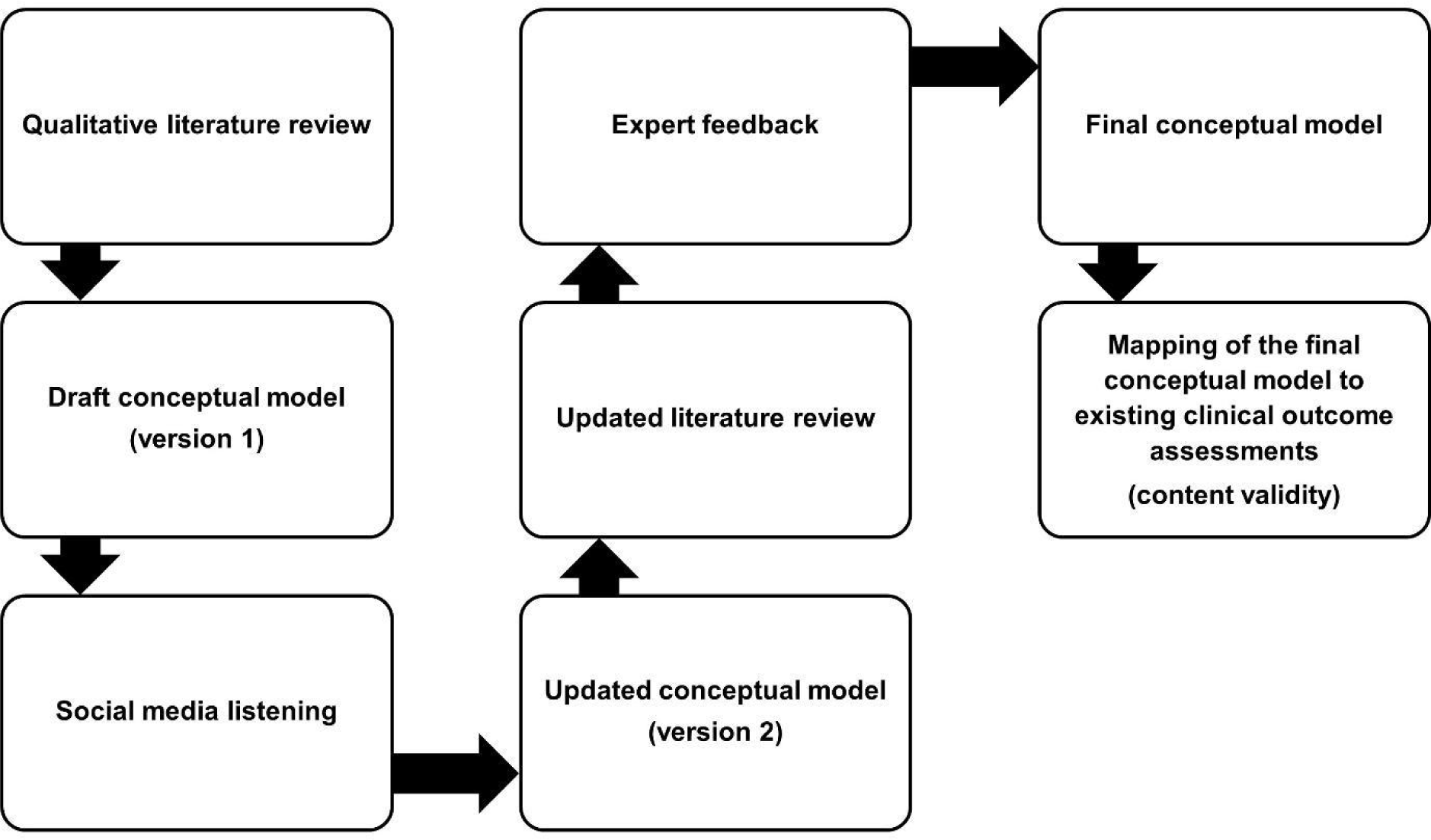
Stepwise approach to conceptual model development. *Note.* Steps in the conceptual model development from the literature search to the confirmation of content validity.

### 2.1. Review of published literature

In March 2021, a two-step targeted literature review of patient-focused research was conducted in PubMed. The aim of this literature review was to understand the patient experience of individuals with SAD, focusing on symptoms and impacts on functioning in daily life. The literature search did not require Institutional Review Board approval. In July 2024, a second literature search was performed to confirm whether any new information was available since the first search.

Step one identified studies that used qualitative interview methods to assess symptoms or functional impacts of having SAD and any previously developed conceptual models of social anxiety. The search strategy for step one was ‘qualitative study’ (All Fields) OR ‘qualitative research’ (All Fields) OR ‘qualitative interview’ (All Fields) AND ‘1000/1/1:2021/3/31[pdat]’ AND ‘social anxiety’ (All Fields) OR ‘social anxiety disorder’ (All Fields) AND ‘1000/1/1:2021/3/31[pdat]’ AND ‘English’ (Filter) AND ‘1000/1/1:2021/3/31[pdat]’ AND ‘English’ (Filter).

Step two identified studies using surveys or questionnaires to determine SAD symptoms and how these impacted individuals with SAD in their daily life. The search strategy for step two was ‘questionnaire’ (All Fields) OR ‘survey’ (All Fields) AND ‘1000/1/1:2021/3/31[pdat]’ AND ‘impact’ (All Fields) OR ‘function’ (All Fields) AND ‘1000/1/1:2021/3/31[pdat]’ AND ‘social anxiety’ (All Fields) OR ‘social anxiety disorder’ (All Fields).

Qualitative research, surveys and questionnaires were included to develop the draft conceptual model. Included publications were written in the English language, with no restrictions on the date of publication. Studies were excluded if topics of the qualitative interviews were not related to symptoms or impacts of having SAD. Full inclusion and exclusion criteria can be found in Table S.1.

### 2.2. SML

SML refers to the systematic use of manual and automated methods that extract relevant insights from social media data for observational studies [25]. It has been shown that SML can be performed following recognized ethical and legal standards to deliver positive results to patients [29]. SML has already been applied in research into mental health conditions [21, 30] such as depression [30, 31], attention deficit hyperactivity disorder, and anxiety [32]; however, most SML studies related to mental health have used an aggregate approach, studying the language and characteristics of large quantities of social media data without delving into specific patient cohorts or validating the presence of a diagnosis. Here we performed a retrospective observational study in a cohort of social media users with evidence of a self-reported SAD diagnosis. This enabled a rigorous analysis of social media data by systematically gathering publicly shared insights. The cohort selection process is described further in section 2.2.2.

Three methodological approaches were employed in SML (described in sections 2.2.3 and 2.2.4):

1. Natural language processing (NLP) analysis for symptom extraction (physical and emotional) and impacts without a priori knowledge of the draft conceptual model based on published literature.
2. NLP analysis with the draft conceptual model to further insights into impacts.
3. Exploratory topic modeling.

#### 2.2.1. Data collection

The data collection and cohort selection process have been described previously by Schmidt et al., 2023 [27]. Social media data were collected from Reddit, a social news website and forum where content, grouped in topic-specific forums known as subreddits, is socially curated and promoted by site members through voting. Reddit member registration is free and anonymous. Registration is required to use the site’s basic features, although no account is required to read the content as it is made publicly available. Institutional Review Board approval for the SML data collection from Reddit was not required as it pertained to public information. Patient consent from Reddit users was not necessitated due to the observational nature of the study, its research objectives and study design [29].

r/SocialAnxiety is a support subreddit about SAD. Like all subreddits, r/SocialAnxiety is moderated and has a set of rules users must follow to keep discussions cordial and on topic. All posts from this subreddit were retrieved using the Pushshift Application Programming Interface (API; Jason Baumgartner) [21]. The Pushshift API was a social media data collection, analysis, and archiving application [21]. It was used for the mining, analysis, and archiving of social media data collected from Reddit. Data through the API application were made available to researchers from 2015 to 2023. [21]. While the Pushshift API is no longer active, the data are still available and can be found at Academic Torrents [33].

The dataset consisted of all the posts on the r/SocialAnxiety subreddit available at the time of download, March 23, 2022. Posts can be of two types:

1. A thread head, which refers to the first post of a new thread within the subreddit. These posts have a title that labels the new thread.
2. A comment, which refers to any post made in an existing thread.

The dataset elements consisted of the post ID, the author, the title of the thread (if a thread head), post text, and a timestamp. The full dataset consisted of 535,544 posts from 118,040 unique users from January 10, 2015, until the date of collection (March 23, 2022).

#### 2.2.2. Cohort selection

A cohort was extracted from all the users on the r/SocialAnxiety subreddit. Eligible Reddit users were aged 13–25 years at the time of posting with reported evidence of clinically relevant social anxiety. A mention of an SAD diagnosis or therapy for SAD was used as a surrogate for clinically confirmed SAD. To be eligible for inclusion, Reddit users had to state their age in a post. The age criteria were based on the median age of onset of SAD at 13 years and focused on a relatively young cohort (up to 25 years of age) before development of comorbidities that may change the phenotype of SAD [34]. In accordance with Reddit’s Terms and Conditions, individuals were required to be at least 13 years of age to sign up and participate on the forum.

Where users reported multiple conditions, it was generally not possible to determine the primary diagnosis from their posts. In traditional SAD studies, participation is usually limited to people for whom SAD is their primary diagnosis [35]. Therefore, to minimize misrepresentation of specific symptoms and impacts unrelated to SAD itself, users were excluded if they reported comorbidities such as substance abuse, and/or if they reported any of the following disorders: generalized anxiety disorder, panic disorder, bipolar disorder, psychotic disorder, schizoaffective disorder, obsessive-compulsive disorder, personality disorder, major depressive disorder, persistent depressive disorder, schizophrenia, or post-traumatic stress disorder.

The cohort selection process is shown in Fig. S.1. For each criterion, regular expressions were used to extract relevant text data followed by manual annotation. Age annotation was done on individual posts, while for all other steps (diagnosis, therapy, and comorbidities), text was chronologically aggregated at the user-level before annotation. The annotation guidelines relevant to the different steps have been published previously [27].

To eliminate errors in single annotated data, the annotation of age, diagnosis, and therapy was double annotated with an unweighted Cohen Kappa of 0.967, 0.948, and 0.876, respectively. The annotation of comorbidities was annotated by one annotator and a random sample of 10% was double annotated to calculate the inter-annotator agreement. In these cases, multi-label annotation was permitted. The unweighted Fleiss’ Kappa was 0.816 and 0.853.

#### 2.2.3. Symptom extraction

##### 2.2.3.1. Physical symptoms

A variety of NLP techniques were used to extract the symptoms from the cohort’s posts. Text was matched to a disease ontology with the commercial tool IQVIA NLP (IQVIA, UK) (formerly known as Linguamatics I2E) to enable the extraction of diseases, disorders, and symptoms from the text [36]. IQVIA NLP’s ontology is derived from Medical Subject Headings, Medical Subject Headings Supplemental Concept Record, and the National Cancer Institute Thesaurus, provides a good coverage of synonyms, and uses morphological variants and dialect variants [36]. Symptom matches within the posts were then manually reviewed and corrected. Any additional new symptom synonyms identified through manual review were searched and newly identified posts annotated. The process was repeated post-by-post in a recursive fashion until reaching terminology saturation, i.e. no new symptom synonyms were identified [37].

##### 2.2.3.2. Emotional symptoms

To complement the results from the ontology-based physical symptom extraction, we devised a simple approach to extract emotional symptoms. Firstly, all mentions of “social anxiety” and its variants in the corpus were masked with a more generic “disease” label. Secondly, part-of-speech tagging was performed for all posts. Part-of-speech tagging is a process in which words in text are marked according to their role as part of speech (noun, verbs, conjunction, adverbs, etc).

Once the text was properly tagged, any sentences with negation were removed and the noun, verbs, adverbs, and adjectives from the remaining text were retained to then lemmatize them. Lemmatization is a process in which different inflected forms of the same word are mapped to one form, for example, run would be the lemma of run, runs, ran, and running.

The generated lemmas were then matched against the WordNet-Affect Lexicon [38], a hand-curated collection of emotion-related words classified as positive, negative, neutral, or ambiguous categorized into 28 subcategories. For this study, the focus was on the negative subcategories: Fear, sadness, dislike, ingratitude, shame, compassion, humility, despair, anxiety, and daze.

#### 2.2.4. Extraction of impacts on the quality of life (QoL)

##### 2.2.4.1. Terminology saturation

Similarly to the extraction of physical symptoms, terminology saturation was used to extract QoL impacts. The only difference was that the seed ontology used was not based on standard ontologies but rather on a set of known QoL impacts associated with SAD based on the published literature. This ontology scope limitation did not prevent the finding of QoL impacts unrelated to the seed terminology.

##### 2.2.4.2. Conceptual topic modeling

To complement the terminology saturation approach, Latent Dirichlet Allocation was used to generate a topic model to classify the posts according to their content. Latent Dirichlet Allocation is a topic modeling algorithm used to discover underlying topics in corpora. It establishes associations between latent topics and tags [39].

The final model was generated after an iterative process of fine-tuning the data pre-processing and model parameters (number of topics, number of iterations, passes). This optimized the statistical measurement of coherence and perplexity, while considering the agreement and confidence score provided by two annotators when labeling the topics in the different iterations. A topic threshold was used to remove automatic labels when the confidence score of the model was low. The final conceptual model was used to classify all posts written by the total cohort and then to calculate the topic prevalence.

### 2.3. Expert review of the conceptual model

Input from experts, including a clinical expert and a patient advocate with experience in SAD, was sought prior to finalization of the conceptual model. The experts reviewed the draft conceptual model, methodology, and conclusions. They noted any missing symptoms or impacts, mapped the topics to the model and raised any concerns with the methodology and limitations of this research. Their feedback was incorporated as part of the iterative process described in section 2.2.4.2 and was reflected in the final conceptual model.

### 2.4. Mapping of the conceptual model to existing clinical outcome assessments (content validity)

Finally, the concepts contained in the conceptual model were mapped by a researcher in a high-level assessment to items in existing clinical outcome assessments commonly used in SAD (Liebowitz Social Anxiety Scale [LSAS] and Brief Social Phobia Scale [BSPS]), to determine the instrument’s content validity [40, 41]. Content validity is the degree to which the content of an instrument, like the conceptual model, is an adequate reflection of the measured construct i.e. concepts for SAD. The LSAS is a 24-item questionnaire commonly used in SAD clinical trials [40]. There is a clinician-reported and patient-reported version, both rated on a 4-point Likert scale, from 0 (none) to 3 (severe) for fear or anxiety and 0 (never) to 3 (usually) for avoidance [42]. The BSPS is a clinician-reported 11-item questionnaire used to measure social phobia symptoms. It includes seven items related to commonly feared or avoided situations, rated from 0 (none) to 4 (extreme fear) for fear and 0 (never) to 4 (always avoidance) for avoidance, and four items related to physiological symptoms, rated from 0 (none) to 4 (extreme) [43].

## 3. Results

### 3.1 Review of published literature

From the two-step targeted literature review, a total of 146 abstracts were identified: 27 results from step one and 119 results from step two.

After screening, 11 studies were included in the analysis (Fig. S.2 and Table S.2); three qualitative studies (*n* = 47) [44-46], one study with qualitative and survey data (*n* = 142) [47], and seven studies with survey and/or questionnaire data (*n* = 3099) [48-54]. The individuals in these studies (*n* = 3288) were aged 15–68 years. The studies included more females (*n* = 1599) than males (*n* = 902).

Three key symptom domains (physical, negative automatic thoughts, and emotions) and two impact domains (social functioning and occupational/educational functioning) were identified from the literature (Table S.3). A draft conceptual model was developed from this data (Fig. S.3).

### 3.2 SML

Out of 118,040 unique users identified on the r/SocialAnxiety subreddit; 945 were eligible for inclusion in the final SML cohort. These users had a total of 28,818 posts. The age distribution for included individuals can be seen in Fig. S.4.

Symptom extraction using NLP without a priori knowledge of the published literature found a cluster of acute symptoms (Fig. 2). A total of 847 posts from 376 users reported at least one of the following symptoms: stuttering, selective mutism, nausea and vomiting, dizziness, cough, blushing, cramps, rapid heart rate, and tremor. These were similar to the physical symptoms included in the draft conceptual model (Fig. S.3). Further, the analysis showed two dimensions: the visibility of symptoms such as stuttering and blushing, as well as strategies to cope with the symptoms (ability to hide symptoms temporarily). Symptoms could also be divided into acute and chronic; acute symptoms may be triggered during social exposure, while chronic symptoms may be displayed outside of social exposure as well. The most frequently reported symptoms were depression (64.4%), suicidal ideation (13.0%), and stuttering and pain (9.3% each). Symptoms reported in fewer than 1% of users, and excluded from Fig. 2, were: amblyopia, apraxia, bulimia, chronic pain, dyslexia, eye pain, fasciculation, flushing, gagging, hallucinations, hyperventilation, irritable heart, lethargy, mania, motion sickness, paresthesia, pruritus, rhinorrhea, seizures, sleepiness, speech disorders, tics, vertigo, aggression, anhedonia, auditory perceptual disorders, blurry vision, dyspepsia, learning disabilities, myalgia, sleep deprivation, sleep wake disorders, thinness, chest pain, dyspnea, hot flashes, and syncope. From the WordNet-Affect Lexicon, the proportions of users from the total cohort of 945 users expressing negative emotions were highest for anxiety (91.7%), fear (86.2%), and sadness (80.5%). The analysis revealed new emotions such as dislike, despair, feeling dazed, compassion, and humility (Fig. 2).

**Fig. 2.**
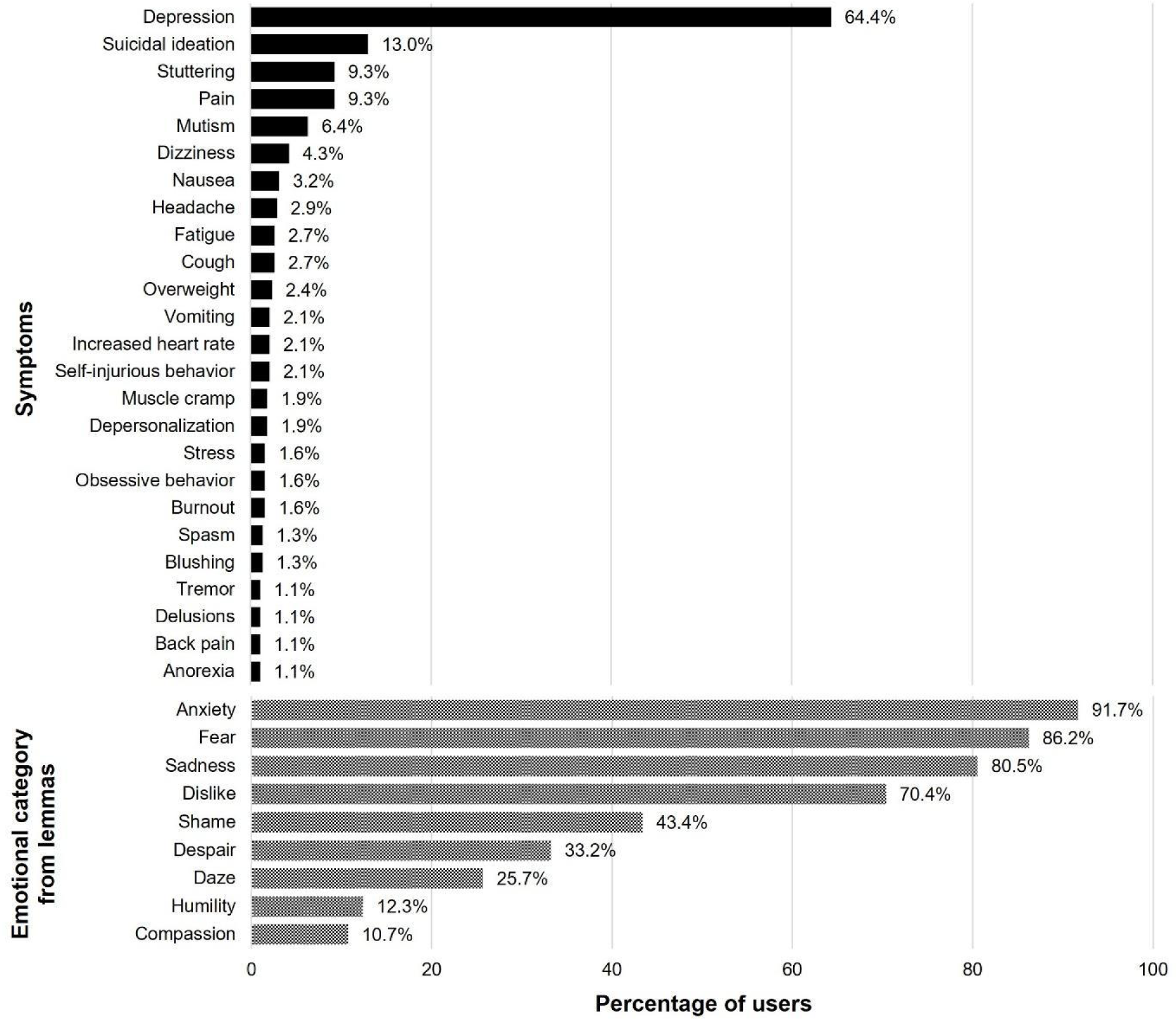
Most frequent symptoms discussed by individuals with SAD on social media. *Note.* Top: Percentage of users from a sample of 376 unique users (847 posts) who mentioned any given symptom. Figure shows symptoms reported in at least 1% of users. Bottom: Percentage of users from the whole cohort of 945 unique users who expressed a negative emotion as categorized by the WordNet-Affect Lexicon. SAD = social anxiety disorder.

The impact of SAD was determined from a group of 93 individuals, representing the number of users from the SML cohort who described at least one impact on daily living (Fig. 3). Similar impacts were identified to those in the literature review, with a focus on social functioning (e.g. socializing [57.8%], starting a relationship [28.9%], making friends [26.5%], and group activities [15.7%]) and occupational or educational functioning impacts (e.g. school participation [30.1%], performing at work [18.1%], and finding a job [10.8%]).

**Fig. 3.**
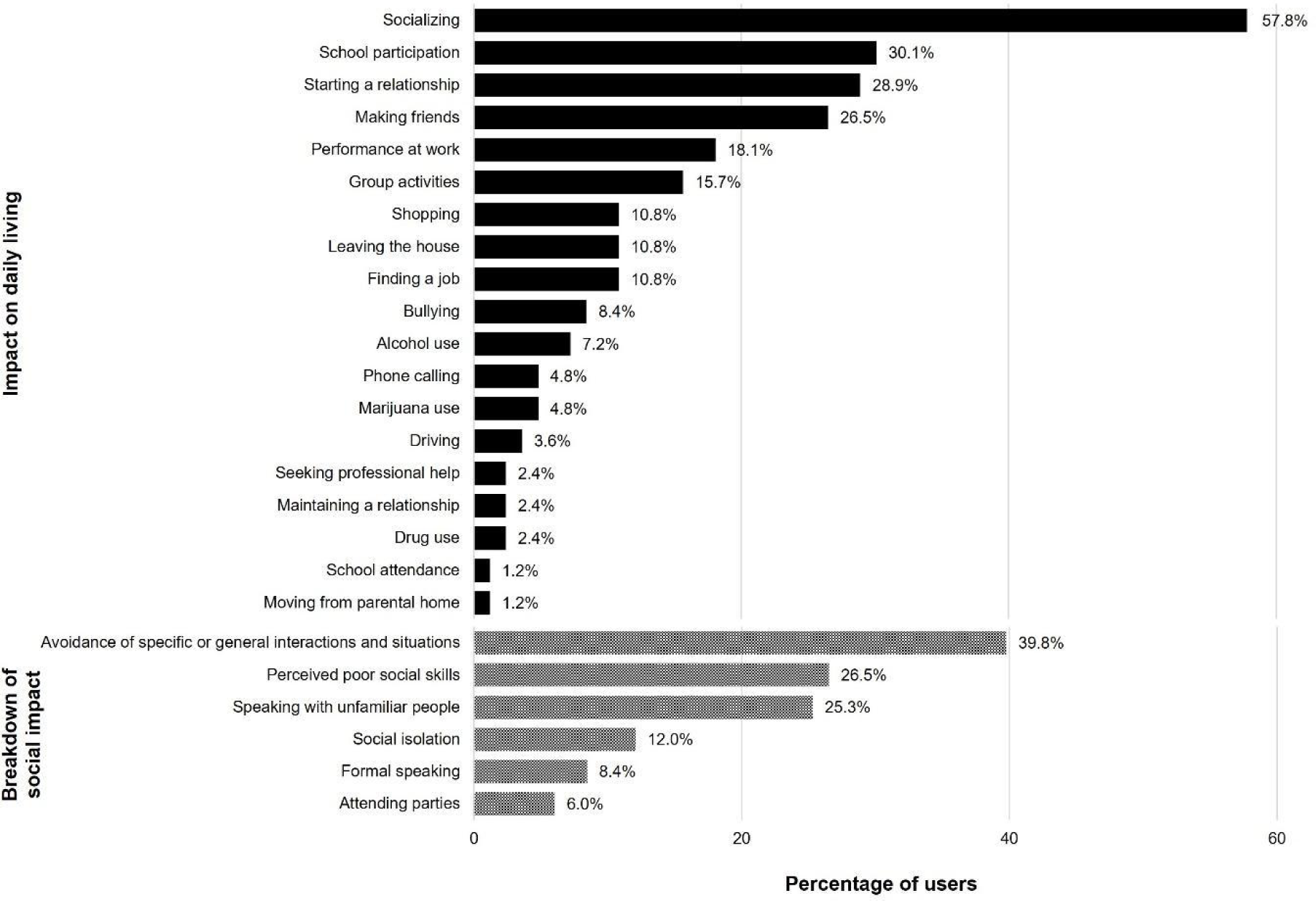
Identified impacts on functioning or daily life discussed by individuals with SAD on social media. *Note.* Top: Percentage of users from a sample of 93 unique users (447 posts) who mentioned any given impact on daily living. Figure shows impact reported in at least 1% of users. Bottom: Breakdown of social impact on daily living into more defined categories and the percentage of users who discussed them. SAD = social anxiety disorder.

Given the high frequency of ‘socializing topics’ discussed online, further investigation of the social impacts of SAD was undertaken. The social functioning concepts within the conceptual model were used as a basis to explore the frequency of posts raising these topics (Fig. 3). The most frequently reported social impacts were avoidance of specific or general interactions and situations (39.8%), perceived poor social skills (26.5%), and speaking with unfamiliar people (25.3%). All concepts in the draft conceptual model were mentioned online, except for “being assertive”.

Further exploratory data analysis approaches were employed with automatic topic modeling to explore the online communities’ SAD symptom and impact presentation. Thirteen discussion topics were identified and mapped to the draft conceptual model (Fig. 4). The five topics most discussed were socialization (88.9%), friends and school (87.3%), feelings about living with SAD (86.6%), family (69.8%), and therapy (68.6%).

**Fig. 4.**
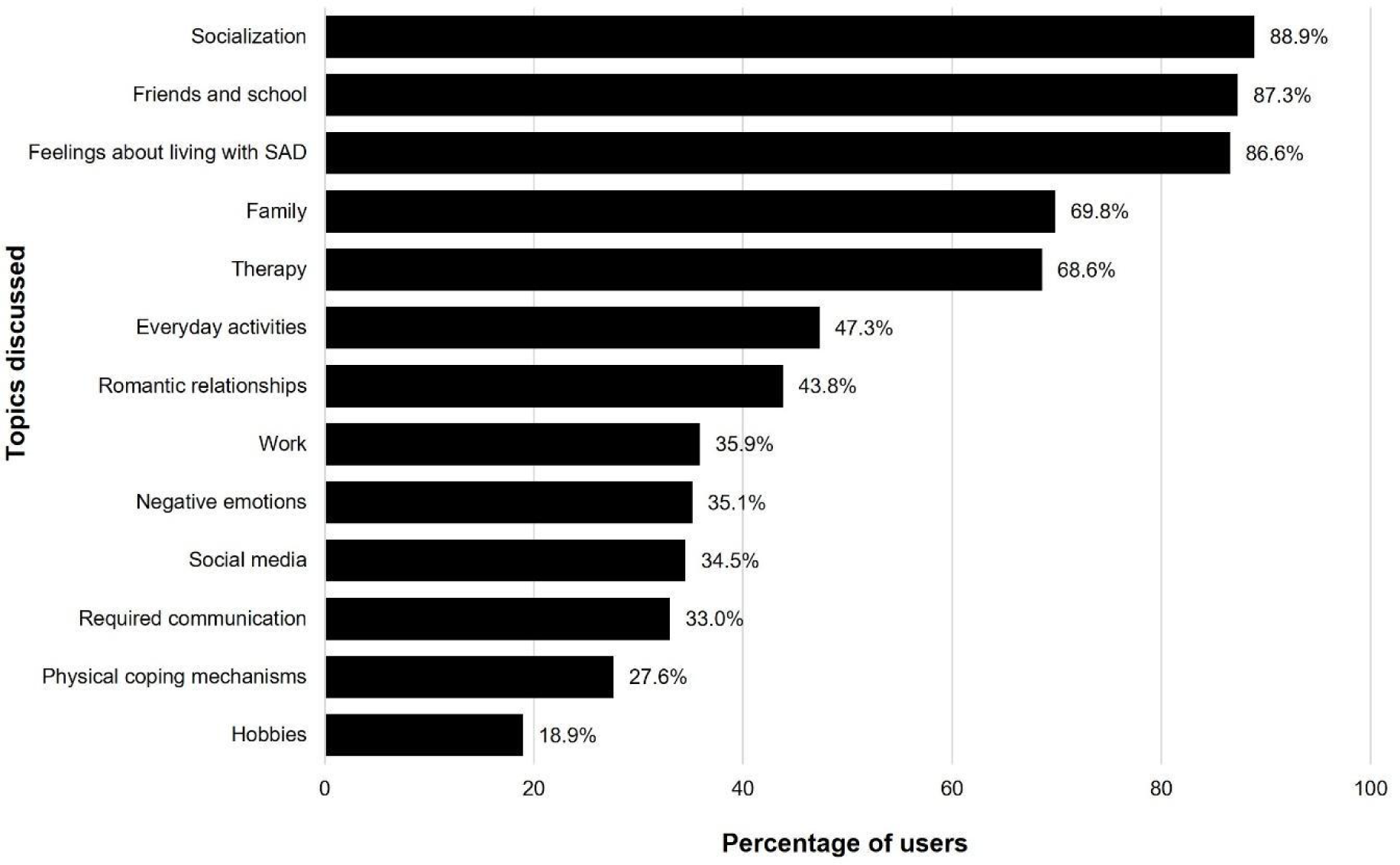
Topics most often discussed by individuals with SAD on social media. *Note.* Figure presents the percentage of users from the cohort of 945 users who discussed a given topic as determined by the topic model. SAD = social anxiety disorder.

Using the draft conceptual model, the overlap of concepts between published literature and social media was explored. There was good convergence in the three key anxiety symptoms included in the draft model, namely physical symptoms, negative automatic thoughts, and emotions, with 20/22 (90.9%) of the specific symptoms reported in both the published literature and SML. The published literature described two unique physical symptoms, namely dissociation and heart palpitations, which were not described on social media.

There was also good overlap in many of the impacts on daily life. For social functioning, 9/10 (90.0%) of the concepts overlapped. Difficulties being assertive was the only impact raised in the published literature that was not captured by social media, indicating that it may not be a commonly used term or concern. Using a threshold of a concept being raised in more than 10% of posts, the social media analysis also found unique physical (pain, e.g. headache; Table S.4), emotional (despair), and social functioning concepts (difficulty leaving the house, starting a romantic relationship, going shopping, having face-to-face informal communication, and difficulty with group activities) which were added to the conceptual model.

The conceptual model was further refined after input from a clinical expert and a patient expert experienced in SAD. The final SAD conceptual model incorporating 13 topics from the published literature and SML is presented in Fig. 5. Researchers mapped the concepts from the conceptual model to the items in existing clinical outcome assessments for SAD (i.e. the LSAS and BSPS scales) to determine the instrument’s content validity. Overall, the mapping assessment revealed good conceptual coverage of the physical symptoms and impacts of social functioning and occupational/educational functioning with the selected SAD scales (Table S.5).

**Fig. 5.**
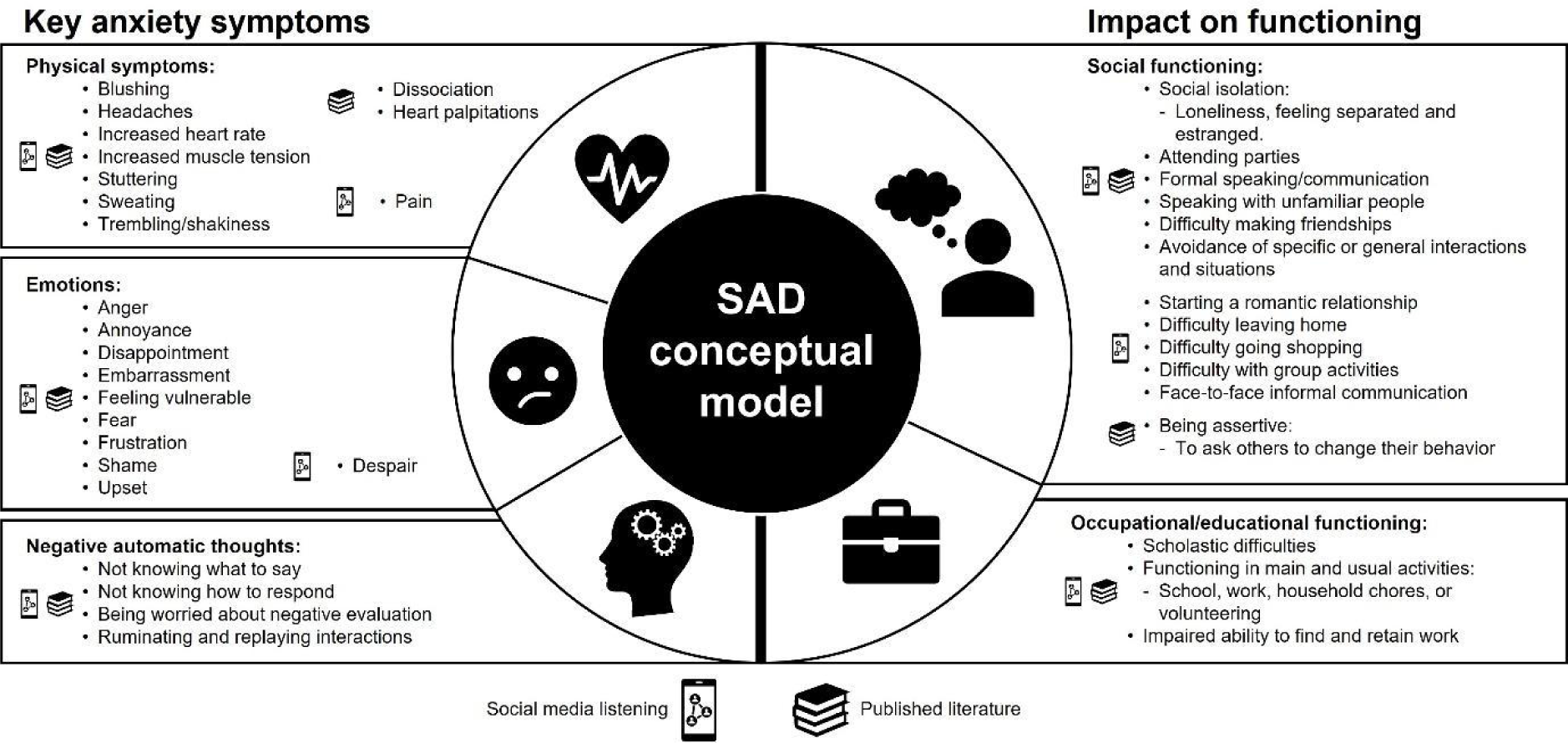
Final conceptual model proposed for SAD. *Note*. Icons of social media listening and/or published literature denote where symptoms and impacts were reported and entered the model. SAD = social anxiety disorder.

## 4. Discussion

These results confirm the importance of using a fit-for-purpose approach when studying each population or disorder. In this case, drawing upon an SML approach rather than requiring participants with SAD to be comfortable with sharing information in an interview or group setting, helped establish a comprehensive conceptual model.

Overall, there is a significant symptom and impact burden for individuals with SAD. The published literature firstly provided an overview of the symptoms and impacts but lacked depth regarding the current lived experience of individuals with SAD due to a paucity of qualitative research. The SML analyses allowed for a deeper evaluation, confirming many of the concepts already in the model and raising new concepts not previously identified in the literature. These results validated the approach of using a listening rather than an elicitation approach, customized to the specific patient population.

Despite strong overlap between the evidence from different methods, there were areas of differentiation, which may be related to the method used. For example, some of the socializing concepts (e.g. starting a relationship) may not be topics that an individual with SAD would bring up directly to a researcher as part of a qualitative study, and they may be more likely to contribute to this discussion online on an anonymous forum or with their therapist. For other topics (e.g. difficulty going shopping), it was less clear why these did not arise in the literature reviewed. Social media represents an optimal approach for supplementing SAD research given that a key concept of the disorder is having difficulty with formal and/or informal communication with strangers.

The final conceptual model can be used to assess the adequacy of existing clinical outcome assessments commonly used in SAD, as done in a high-level approach here with the BSPS and LSAS. While many physical symptoms were mapped to BSPS, and social functioning and occupational/educational functioning items mapped to both LSAS and BSPS, there were concepts included in the model within these domains that were not directly covered by items in either scale (e.g., increased heart rate, difficulty leaving the house, and difficulty making friendships). Furthermore, negative automatic thoughts were not mapped to either the LSAS or BSPS. In the context of clinical studies, one approach to ensure comprehensive assessment of symptoms and impacts based on the new conceptual model is to supplement well-established clinical outcome measures with alternative scales that cover additional parts of the model such as the broader emotional impact of SAD and the negative automatic thoughts, to ensure adequate conceptual coverage of all domains.

Limitations of the study should be considered, such as that the review of the published literature was not systematic and hence publications of relevance may have been missed. Additionally, due to the methodology of manually reviewing the symptoms experienced, some uncommon symptoms may have been overlooked. The literature and social media sources did not match in terms of demographics of the populations included. The SML analysis was limited to individuals reporting an age of 13–25 years and the data may not be generalizable to an older population. This was accepted as the focus of this study was SAD in early life, before the development of major comorbidities. There may be differences in topics discussed by adolescents and adults with SAD included in this study (e.g. friends and school, and the ability to talk about complex emotions). However, the focus was to develop a conceptual model that would be viable across an adolescent/young adult population. Additionally, data could include recall or reporting bias (when individuals do not remember specifics of past events, selectively disclose information, or omit details) [55], or post-event processing (when an individual’s repeated consideration and/or potential reconstruction of events leads to incorrect reporting) [56].

Social media data are inherently limited by what users choose to disclose. It is very likely that many users matched the selection criteria but were excluded from the cohort because they did not explicitly report their characteristics, for example, they did not report their age. An especially difficult area in this regard is the exclusion of individuals who do mention common comorbid disorders, which in classic psychiatric studies is often addressed by requiring the disease of interest (here, SAD) to be the primary disorder. Due to the nature of SML, no hierarchy can be established between different diagnoses mentioned, resulting in an inevitable mismatch between inclusion criteria in traditional human subject research and SML methods. Likewise, it is possible that a user had another mental disorder diagnosis and/or substance abuse disorder and did not disclose this. It was not possible to verify diagnoses nor was there a way to verify the truthfulness of the user statements. Finally, a threshold of 10% of mentions was arbitrarily chosen for the inclusion of symptoms and impacts retrieved from SML as there are no existing gold standards for this methodology, however this threshold has been used in other studies [24].

## 5. Conclusions

There is a significant symptom and impact burden for individuals living with SAD. This study provides an in-depth understanding of the lived patient experience, integrating complementary sources of published literature and social media research. It also highlights the value of social media research when it is not optimal to use traditional qualitative research methods, such as interviews or focus groups for specific populations. The final conceptual model will be valuable for the understanding of the disorder and for optimizing research outcomes for SAD.

## Data Availability

The dataset consists of Reddit social media data that is public for anyone and can also be downloaded at Academic Torrents: https://academictorrents.com/details/7c0645c94321311bb05bd879ddee4d0eba08aaee

## Acknowledgments

We thank Chantal Dolan and Kathryn Fisher from CMD Consulting, Inc. who performed the targeted literature review, and Andrea de Kock, PhD, of Nucleus Global, an Inizio company, for providing medical writing assistance for the manuscript in accordance with Good Publication Practice (GPP) 2022 guidelines (https://www.ismpp.org/gpp-2022). This study was funded by F. Hoffmann-La Roche Ltd, Basel Switzerland.

## Glossary

API: Application Programming Interface
BSPS: Brief Social Phobia Scale
CBT: Cognitive Behavioral Therapy
FDA: US Food and Drug Administration
LSAS: Liebowitz Social Anxiety Scale
NLP: Natural language processing
QoL: quality of life
SAD: Social anxiety disorder
SML: Social media listening
SoC: standard of care
SSRI: Selective serotonin reuptake inhibitors

