## Supplementary Material for "The lived experience of social anxiety disorder: A conceptual model based on published literature and social media listening"

This supplement has been provided by the authors to give readers additional information about the study.

**Contents:**


**Fig. S.1.** Process of cohort selection for SML

**Fig. S.2.** PRISMA flow diagram for the literature review in SAD

**Fig. S.3.** Draft conceptual model based on published literature

**Fig. S.4.** Age distribution of the SML cohort

**Table S.1.** Inclusion and exclusion criteria for the targeted literature review

**Table S.2.** Overview of studies in individuals with social anxiety that contributed to the conceptual model

**Table S.3.** Key symptom and impact domains identified in individuals with SAD: Selected quotes extracted from the published literature

**Table S.4.** Physical pain symptoms identified in individuals with SAD: Selected quotes were extracted and adapted from social media to ensure anonymity

**Table S.5.** Researcher manual mapping of the concepts from the SAD conceptual model to items in the LSAS and BSPS

Fig. S.1. Process of cohort selection for SML

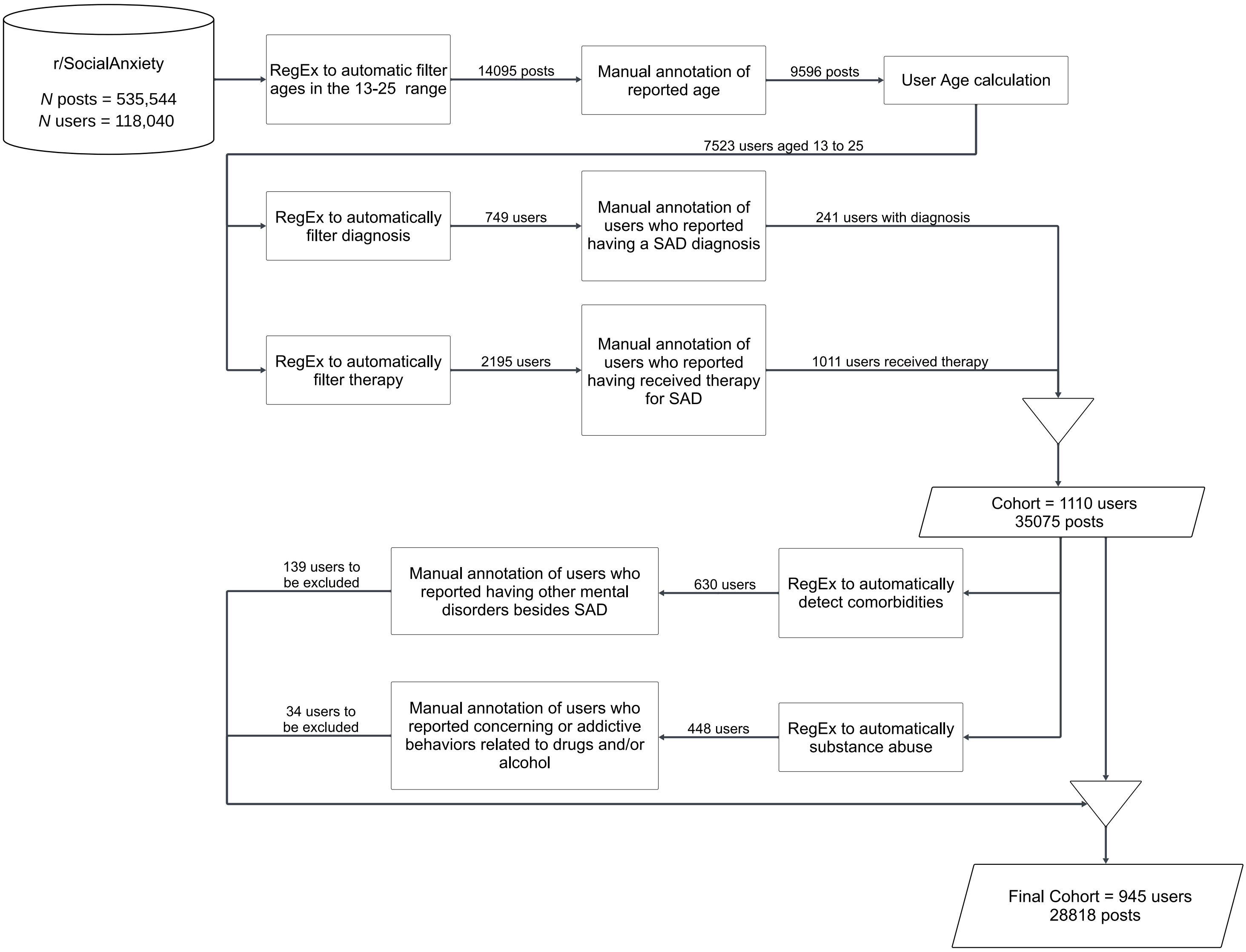


*Note.* For diagnosis, therapy, comorbidities and substance abuse steps, text was chronologically aggregated at the user-level before annotation. Age annotation was done on individual posts, and user could be associated with multiple ages over time on r/SocialAnxiety. A reference age was estimated for each user. All age annotations were used to calculate their birth year. When a user had conflicting age annotations, the median birth-year value of the set was used. For the analysis, the reference age of the user was the year the post was written minus the estimated birth year. The user activity duration had a long-tail distribution in which the median was 16 days, the mean was 181 days and the standard deviation was 323 days. This reference age is a fair approximation of the users' ages for these analyses.

RegEx = machine learning regular expressions tool, SAD = social anxiety disorder, SML = social media listening.

Fig. S.2. PRISMA flow diagram for the literature review in SAD

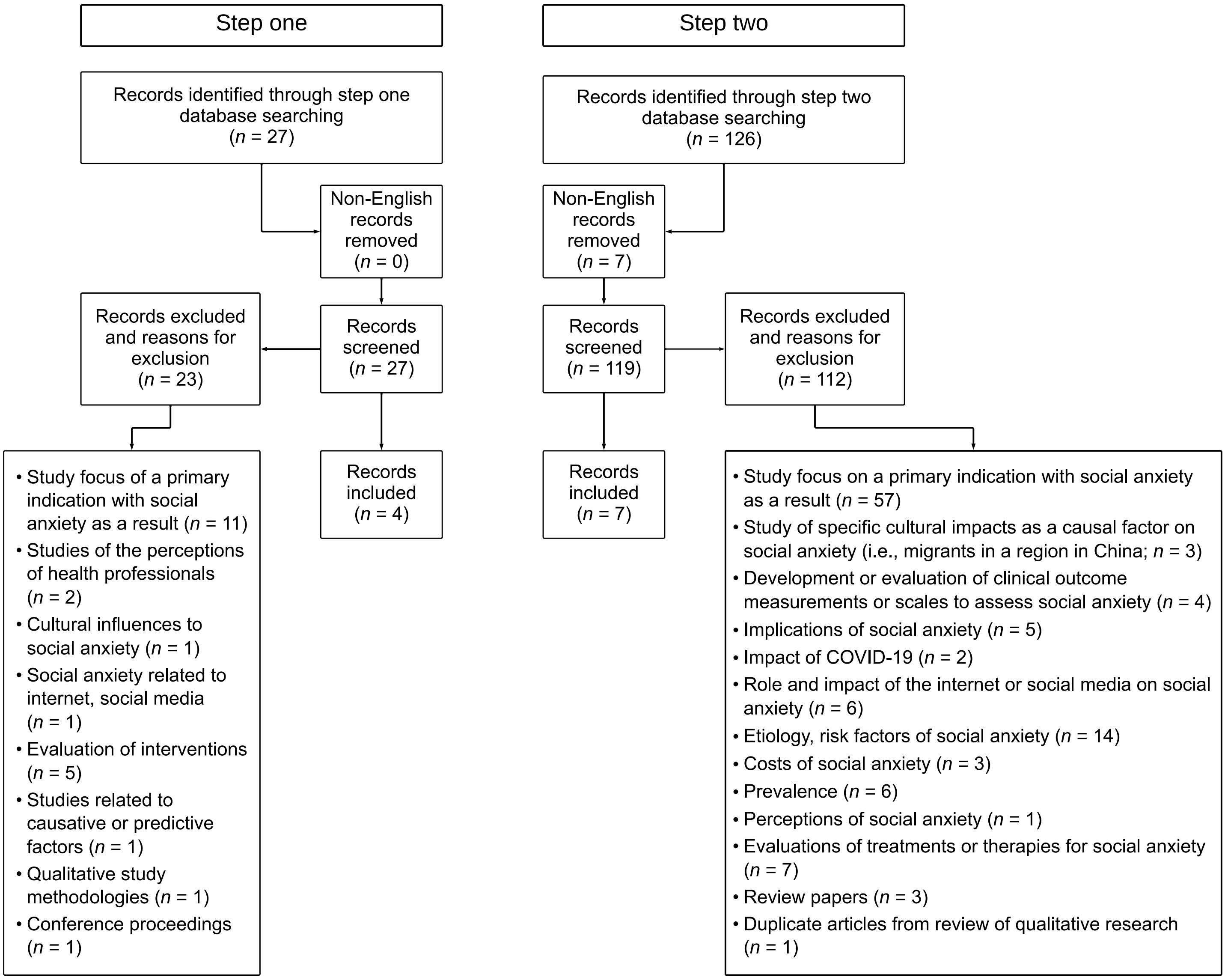


COVID-19 = coronavirus disease 2019, PRISMA = Preferred Reporting Items for Systematic reviews and Meta-Analyses, SAD = Social anxiety disorder.

Fig. S.3. Draft conceptual model based on published literature

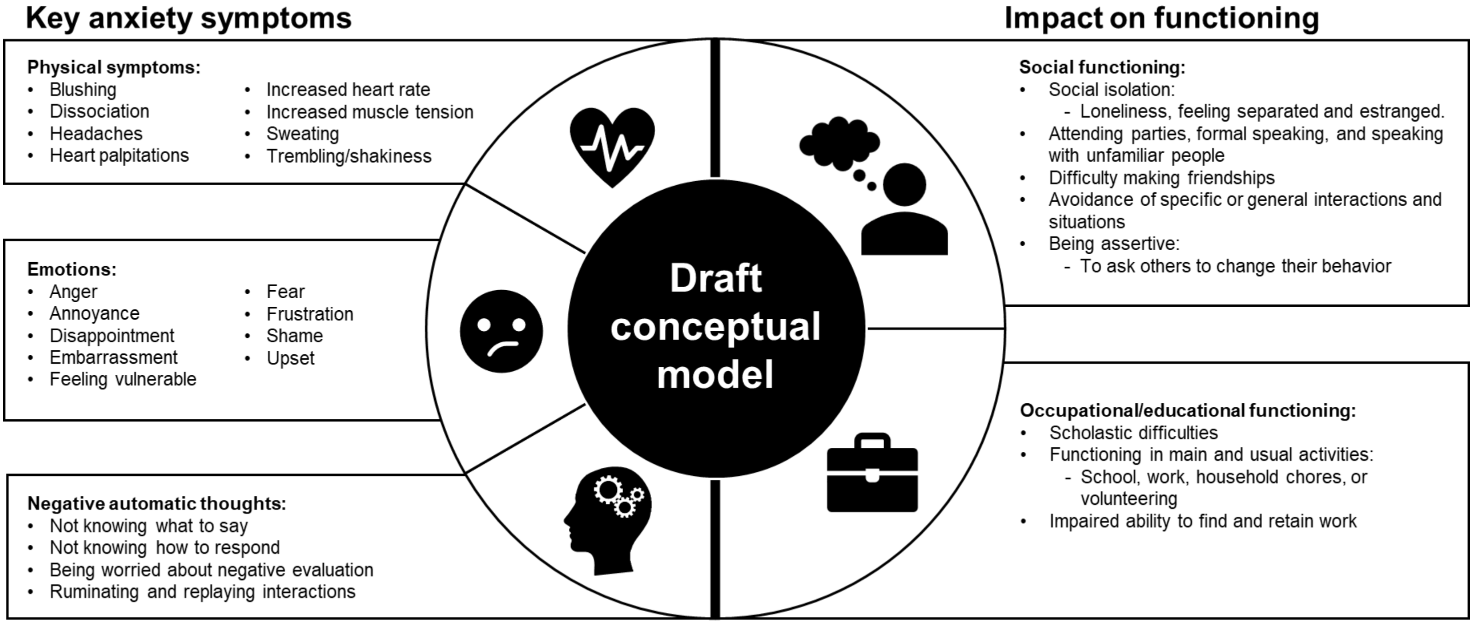

Fig. S.4. Age distribution of the SML cohort

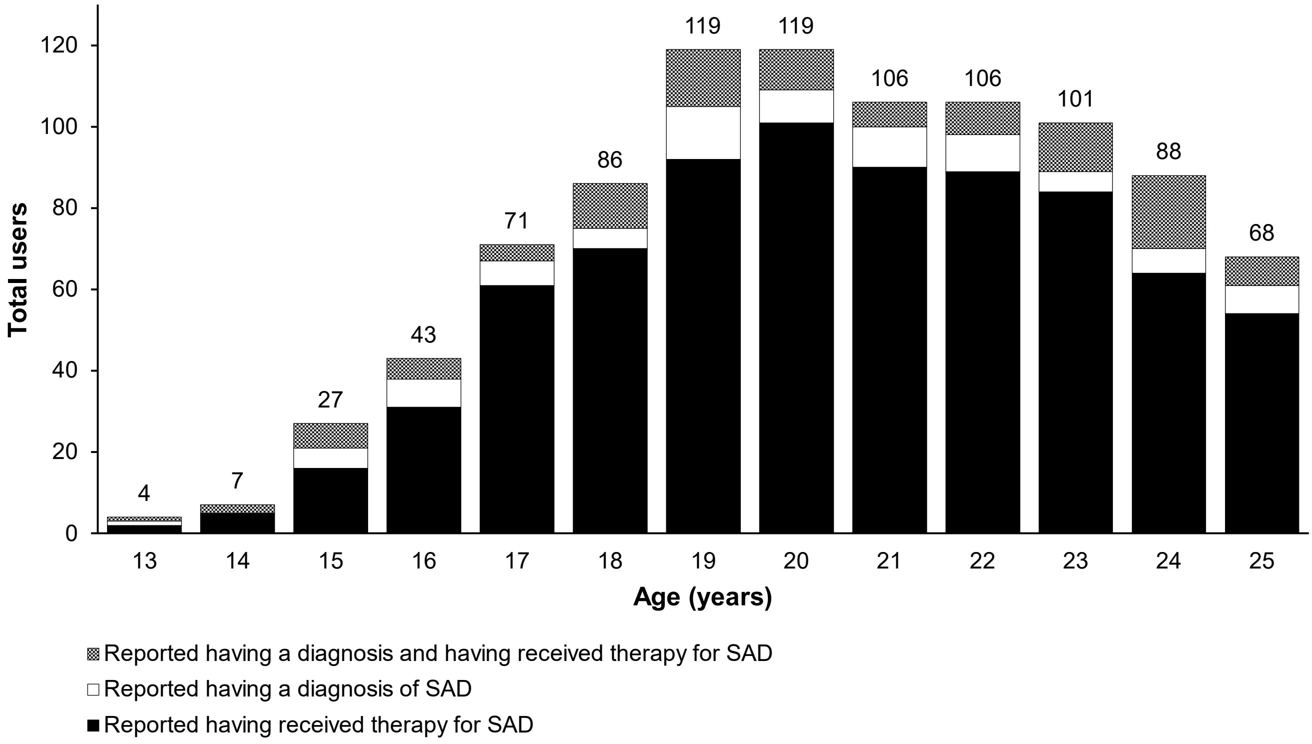


*Note.* Figure presents the number of social media users reporting diagnosis of SAD or therapy for SAD.

SAD = social anxiety disorder, SML = social media listening.

Table S.1. Inclusion and exclusion criteria for the targeted literature review

| **Step one: Literature review of qualitative research studies in individuals with SAD** | |
|---|---|
| **Inclusion** | **Exclusion** |
| • Study participants: Studies of individuals with a diagnosis of SAD or seeking treatment for social anxiety | • Review articles without study data<br>• Studies in people with a primary diagnosis or indication other than social anxiety |
| • Studies: Qualitative studies published as a full-text original article in English that include qualitative study data and/or use a survey or questionnaire to identify symptoms or functional impacts of social anxiety<br>• No restriction on date of publication | • Studies published in a non-English language<br>• Studies of etiology of social anxiety<br>• Studies that did not assess symptoms caused by social anxiety or functional impacts as a result of having social anxiety<br>• Studies that looked at social anxiety as a result or complication of a primary indication |
| **Step two: Literature review of studies using surveys or questionnaires to assess the functional impacts of having SAD** | |
| **Inclusion** | **Exclusion** |
| • Study participants: Studies in a general population or random sample | • Review articles without study data<br>• Studies in people with a primary diagnosis or indication other than social anxiety |
| • Studies: Studies published as a full-text original article in English language using a survey or questionnaire to assess functional impacts or impairment as a result of social anxiety<br>• No restriction on date of publication | • Studies published in a non-English language<br>• Studies of etiology of social anxiety<br>• Studies that looked at social anxiety as a result or complication of a primary indication<br>• Studies evaluating treatments or therapies for social anxiety<br>• Studies of the prevalence of social anxiety<br>• Studies evaluating the role or impact of the internet or social media on social anxiety<br>• Studies evaluating costs of social anxiety<br>• Studies assessing implications of social anxiety |

Table S.2. Overview of studies in individuals with social anxiety that contributed to the conceptual model

| Reference | Demographics | *N* | Data collection method |
|---|---|---|---|
| **Qualitative studies** | | | |
| **Himle (2020) [44]** | • Mean (SD) age of 44 (11.7) years<br>• 8 males (67%), 4 females (33%)<br>• 6 with mild social anxiety (50%)<br>• 6 with severe social anxiety (50%) assessed with the Liebowitz Social Anxiety Scale<br>• Unemployed individuals<br>• US | 12 | • Semi-structured qualitative interviews conducted individually |
| **Hjeltnes (2016) [45]** | • Aged 19–25 years<br>• Mean (SD) age of 23 (1.4) years<br>• 14 males, 15 females<br>• Seeking treatment for symptoms of severe shyness or SAD<br>• Norway | 29 | • Semi-structured qualitative interviews conducted individually |
| **McEvoy (2016) [46]** | • Aged 26–55 years<br>• 4 males (67%), 2 females (33%)<br>• Self-referred to CBT for SAD or diagnosed with SAD<br>• Ireland | 6 | • Semi-structured qualitative interviews conducted individually |
| **Qualitative study including survey data** | | | |
| **Boyle (2019) [47]** | • Aged 16–68 years<br>• 39 males (27%), 103 females (73%)<br>• Participants diagnosed or self-diagnosed with social anxiety<br>• UK | 142 | • Cross-sectional survey that allowed detailed response on thoughts, feelings, and socio-spatial surroundings (*n* = 120)<br>• Synchronous online, semi-structured, qualitative interviews (*n* = 19)<br>• Telephone interviews (*n* = 3) |
| **Studies with surveys and/or questionnaires** | | | |
| **Aderka (2012) [48]** | • Aged 18–60 years<br>• Mean (SD) age of 35 (10.1) years<br>• 106 males (49%), 110 females (51%)<br>• All participants had a diagnosis of SAD according to DSM-IV criteria<br>• Israel | 216 | • Cross-sectional survey<br>• Participants completed a battery of self-reported questionnaires: Mini International Neuropsychiatric Interview, LSAS, Montgomery and Asberg Depression Rating Scale, and Sheehan Disabilities Scale |
| **Arditte Hall (2020) [49]** | • Aged 19–64 years<br>• Mean (SD) age of 36 (22.6) years<br>• 45 males (48%), 48 females (52%)<br>• US | 93 | • Cross-sectional survey<br>• Participants completed online questionnaires including vignettes that participants had to rate by forecasting a series of affective and empathic emotions<br>• Participants also completed multiple-choice questions to validate the responses<br>• SAD was assessed using the 20-item Social Anxiety Interaction Inventory |
| **Barnett (2021) [50]** | • Aged 18–30 years<br>• Mean (SD) age of 21 (2.2) years<br>• 233 males (29%), 580 females (71%)<br>• Students recruited through university website<br>• US | 813 | • Cross-sectional survey<br>• SAD was assessed using the Social Interaction Anxiety Scale and Social Phobia Scale |
| **Maddox (2015) [51]** | • Aged 16–45 years<br>• SAD (*n* = 26)<br>• Mean (SD) age of 26 (7.1) years<br>• 13 males (50%), 13 females (50%)<br>• SAD met diagnostic criteria as assessed by Anxiety Disorders Interview Schedule<br>• US | 79<br><br>(*n* = 26 SAD) | • Cross-sectional survey<br>• Participants completed a battery of self-reported questionnaires: Social Responsiveness Scale-2, Brief Fear of Negative Evaluation Scale<br>• Participants also completed the SAD section of the Anxiety Disorders Interview Schedule for DSM-IV: Social Phobia Module. Participants were also asked about when social anxiety symptoms were most intense and impairing |
| **Russell (2012) [52]** | • Aged 16–60 years<br>• 293 (36.5%) were included in age range of 16–20 years<br>• 358 (55.8%) were included in age range of 21–30 years | 787 | • Cross-sectional survey<br>• Explored students' views on impact of social anxiety on well-being and learning<br>• Invited students who experienced issues relating to shyness, embarrassment, and anxiety in public |

| | | | |
|---|---|---|---|
| | • UK | | • SAD was assessed using the three-item Mini-Social Phobia Inventory scale |
| **Stein (2000) [53]** | • Aged 15–64 years<br>• Social phobia in lifetime (*n* = 1116)<br>• 440 males (39%), 676 females (61%)<br>• Social phobia in last year (*n* = 566)<br>• 219 males (39%), 347 females (61%)<br>• Canada | 8000<br><br>(*n* = 1116 social phobia) | • Cross-sectional survey<br>• The Ontario Health Survey Mental Health Supplement |
| **Tolman (2009) [54]** | • Aged 25–54 years<br>• Female (100%)<br>• All received welfare<br>• SAD (*n* = 48)<br>• US | 753<br><br>(*n* = 48 SAD) | • Cross-sectional survey<br>• Participants who received welfare (1997), re-interviewed in 1998, 1999, and 2000<br>• SAD was assessed using the WHO Composite International Diagnostic Interview Short-Form. |

CBT = cognitive behavioral therapy, DSM-IV = Diagnostic and Statistical Manual of Mental Disorders, fourth edition, LSAS = Liebowitz Social Anxiety Scale, SAD = social anxiety disorder, SD = standard deviation, WHO = World Health Organization.

Table S.3. Key symptom and impact domains identified in individuals with SAD: Selected quotes extracted from the published literature

| **Symptom** | **Quote** |
|---|---|
| **Physical symptoms**<br><br>Blushing, dissociation, headaches, heart palpitations, increased heart rate, increased muscle tension, sweating, trembling/shaky | • *"I sweat and blush, shake, stutter and it's like my anxiety is just oozing out for everyone to see." [47]*<br>• *"It's kind of an experience of not having control at all that is so unpleasant that it's… there's not control. When I have those bad days and feel shaky, then I can feel that I react to everything." [45]* |
| **Emotional symptoms**<br><br>Anger, annoyance, disappointment, embarrassment, feeling vulnerable, fear, frustration, shame, upset | • *"I get angry and frustrated with myself…and it feels like I'm terrified inside." [45]*<br>• *"I get disappointed in myself…try to hide that I feel insecure." [45]*<br>• *"I was so disappointed. I was really disappointed but I ain't say nothing." [44]* |
| **Negative automatic thoughts**<br><br>Knowing what to say, how to respond, worries about negative evaluation, ruminating and replaying interactions | • *"…[I] sort of analyze everything I did, analyze all the reactions, the facial expressions of everyone in the situation, everything I think that they were thinking. When I think and think about it, everything become much worse." [45]*<br>• *"At the end of the day I go over in my head every last detail of what I did, what I said, how I said it, what my face looked like, did I make enough eye contact and how were people reacting to me? I'm constantly questioning myself." [47]*<br>• *"Feeling stupid or saying the wrong thing." [44]* |
| **Social functioning**<br><br>• Social isolation – loneliness, feeling separated, estrangement<br>• Attending parties<br>• Formal speaking<br>• Speaking with unfamiliar people<br>• Difficulty making friendships<br>• Avoidance of specific or general interactions and situations<br>• Being assertive to ask others to change their behavior | • *"I actually do feel alone a lot. I feel I cannot reach out to people or friends who understand how it is like for me. I feel very alone in thinking about how I should do things, or I feel that when I finally have an idea about what I should do, then it's not quite what they're asking for, or it's completely wrong…" [45]*<br>• *"It's not that common that I get invited to things anymore. Because I've always said no to social gatherings and things, I do not get invited anymore. It gets pretty lonely then. Much isolation, and it's hard to break that circle, because I feel [that] I cannot expose myself to these social things either, because I kind of … when I do not expose myself, then I do not get any practice, no improvement. And then it just gets worse and worse to get back into it." [45]* |
| **Occupational functioning**<br><br>• Scholastic difficulties<br>• Functioning in main and usual activities (school, work, household chores, or volunteering)<br>• Impaired ability to find and retain work | • *"[I want] to be able to enjoy my work. And…I want to dare to speak up without thinking: "Ok, then I might have to say something in a meeting." Just those kinds of things. And then there is something about not having to experience all that discomfort in my everyday life…just to go out shopping, and stand in a line, just be in a room and situations where there are other people." [45]*<br>• *"I feel that it has hindered me from participating fully and wholly in life." [45]*<br>• *"Say I'm going to the shops; I'll take a longer, quieter route." [47]* |

Table S.4. Physical pain symptoms identified in individuals with SAD: Selected quotes were extracted and adapted from social media to ensure anonymity

| Symptom | Quote |
|---|---|
| **Pain**<br><br>Exhaustion, physical pain | • *"I simply can't manage it. I find myself avoiding classes because being around others overwhelms me and leaves me drained for days. It kills my drive to complete my homework when I know I'll face more stress and pain…"*<br>• *"I completely understand. Sometimes I just want to scream as loudly as possible because it's the only way to alleviate the pain…"* |
| **Chest pain**<br><br>Chest pain, pain in ribs | • *"…Attending any social gathering triggered panic attacks (shaking, sweating, dizziness, difficulty breathing, chest pain)…"*<br>• *"For the past few months, I've been experiencing persistent pain in my ribs. It's not a sharp pain, but it feels like there's a constant pressure on my ribs most of the time."*<br>• *"When I'm not anxious, my resting heart rate is around 60. But when anxiety kicks in, it skyrockets to the 100s. My chest always feels like it's experiencing a heart attack, and it hurts constantly. My doctor mentioned that the pain is due to my anxiety, so it seems there's not much that can be done about it…"* |
| **Stomach pain** | • *"…I was short of breath, and after giving my introduction, my belly hurt so bad. It took some time to disappear…"*<br>• *"Each anxious thought feels like a stab, and I can't ignore the pain in my gut. I don't understand why life has been so painful for me, with so much burden on my shoulders…"*<br>• *"I've experienced stomach aches for as long as I can remember. It's not the intense, panicky kind of pain, but rather a constant, dull discomfort that feels like a knot in my gut all the time…"* |
| **Muscle pain** | • *"My chiropractor advised me to regularly pause and consciously relax my neck and shoulders. Doing this a few times a day can help interrupt the cycle of constant muscle tension and spasms. The more you practice it, the easier it becomes for your body to naturally respond better on its own…"* |
| **Headache** | • *"My symptoms are primarily physical: a pounding heart, tripping over my words, the impending sensation I'm about to cry, headaches, and dizziness…"*<br>• *"After coming home with headaches, I end up lying down for hours, overthinking everything and becoming anxious about what lies ahead."*<br>• *"I know I'm going to struggle with sleep tonight; I'm already feeling a headache coming on and experiencing intense anxiety."*<br>• *"I start by feeling lightheaded, then I become mentally overwhelmed, leading to tears or a headache…"*<br>• *"My previous job in retail caused me so much anxiety that I ended up with severe migraines, taking days to recover. I spent days in bed with the worst headaches I've ever had, vomiting more than five times a day."* |

Table S.5. Researcher manual mapping of the concepts from the SAD conceptual model to items in the LSAS and BSPS

| **SAD conceptual model** | | **Mapped to LSAS** | **Mapped to BSPS** |
|---|---|---|---|
| **Key anxiety symptoms** | *Physical symptoms* | | • Heart palpitations (palpitations)<br>• Sweating (sweating)<br>• Blushing (blushing)<br>• Trembling (trembling) |
| | *Emotions* | | • Embarrassment (being embarrassed or humiliated) |
| | *Negative automatic thoughts* | • *Not assessed in LSAS* | • *Not assessed in BSPS* |
| **Impact on functioning** | *Social functioning* | • Attending parties (going to a party)<br>• Formal speaking or communication (talking to people in authority, acting, performing or giving a talk in front of an audience, speaking up at a meeting, and/or giving a report to a group)<br>• Speaking with unfamiliar people (calling someone you don't know very well, talking with people you don't know very well, and/or meeting strangers)<br>• Being assertive or asking others to change their behavior (expressing a disagreement or disapproval to people you don't know very well)<br>• Starting a romantic relationship (trying to pick up someone)<br>• Difficulty going shopping (returning goods to a store, and/or resisting a high-pressure salesperson)<br>• Difficulty with group activities (participating in small groups)<br>• Face-to-face informal communication (participating in small groups) | • Formal speaking or communication (speaking in public of in front of others)<br>• Speaking with unfamiliar people (talking to strangers)<br>• Attending parties (social gatherings)<br>• Difficulty with group activities (social gatherings) |
| | *Occupational functioning* | • Functioning in main and usual activities e.g. school, work, household chores, and/or volunteering (working while being observed, writing while being observed, speaking up at a meeting, and/or taking a test) | |

*Note*. Exact wording from the items in the LSAS and BSPS are included in brackets.

BSPS = Brief Social Phobia Scale, LSAS = Liebowitz Social Anxiety Scale, SAD = social anxiety disorder.
